# How Does ChatGPT Perform on the Medical Licensing Exams? The Implications of Large Language Models for Medical Education and Knowledge Assessment

**DOI:** 10.1101/2022.12.23.22283901

**Authors:** Aidan Gilson, Conrad Safranek, Thomas Huang, Vimig Socrates, Ling Chi, R. Andrew Taylor, David Chartash

**Affiliations:** Section for Biomedical Informatics and Data Science, Yale University School of Medicine; Department of Emergency Medicine, Yale School of Medicine; Program of Computational Biology and Bioinformatics, Yale University; School of Medicine, University College Dublin - National University of Ireland, Dublin

**Keywords:** Natural Language Processing, MedQA, GPT, Medical Education, Chatbots, Artificial Intelligence, Education Technology

## Abstract

**Background:** ChatGPT is a 175 billion parameter natural language processing model which can generate conversation style responses to user input.

**Objective:** To evaluate the performance of ChatGPT on questions within the scope of United States Medical Licensing Examination (USMLE) Step 1 and Step 2 exams, as well as analyze responses for user interpretability.

**Methods:** We used two novel sets of multiple choice questions to evaluate ChatGPT’s performance, each with questions pertaining to Step 1 and Step 2. The first was derived from AMBOSS, a commonly used question bank for medical students, which also provides statistics on question difficulty and the performance on an exam relative to the userbase. The second, was the National Board of Medical Examiners (NBME) Free 120-question exams. After prompting ChatGPT with each question, ChatGPT’s selected answer was recorded, and the text output evaluated across three qualitative metrics: logical justification of the answer selected, presence of information internal to the question, and presence of information external to the question.

**Results:** On the four datasets, AMBOSS-Step1, AMBOSS-Step2, NBME-Free-Step1, and NBMEFree-Step2, ChatGPT achieved accuracies of 44%, 42%, 64.4%, and 57.8%. The model demonstrated a significant decrease in performance as question difficulty increased (P=.012) within the AMBOSS-Step1 dataset. We found logical justification for ChatGPT’s answer selection was present in 100% of outputs. Internal information to the question was present in *>*90% of all questions. The presence of information external to the question was respectively 54.5% and 27% lower for incorrect relative to correct answers on the NBME-Free-Step1 and NBME-Free-Step2 datasets (P<=.001).

**Conclusion:** ChatGPT marks a significant improvement in natural language processing models on the tasks of medical question answering. By performing at greater than 60% threshold on the NBME-FreeStep-1 dataset we show that the model is comparable to a third year medical student. Additionally, due to the dialogic nature of the response to questions, we demonstrate ChatGPT’s ability to provide reasoning and informational context across the majority of answers. These facts taken together make a compelling case for the potential applications of ChatGPT as a medical education tool.

## INTRODUCTION

ChatGPT, or Chat Generative Pre-trained Transformer, is a 175 billion parameter natural language processing (NLP) model that uses deep learning algorithms trained on vast amounts of data to generate human-like responses to user prompts. ^1^ As a general purpose dialogic agent, ChatGPT is designed to be able to respond to a wide range of topics, potentially making it a useful tool for customer service, chatbots, and a host of other applications. Since its release, it has garnered significant press for both seemingly incredible feats like automated generation of responses in the style of Shakespearean sonnets while also failing to answer simple mathematical questions. ^2–4^

Within the medical domain, language models have been investigated as tools for personalized patient interaction ^5^ and consumer health education. ^6^ While demonstrating potential, these models have had limited success in areas testing clinical knowledge through generative questionanswering (QA) tasks. ^7^,^8^ ChatGPT builds on OpenAI’s previous GPT-3.5 language models with the addition of both supervised and reinforcement learning techniques. ^9^ ChatGPT could represent the first in a new line of models which may better represent the combination of clinical knowledge and dialogic interaction. Although it is not an information retrieval tool like Google Scholar, ^10^ UpToDate, ^11^ DynaMed, ^12^ or PubMed, ^13^ its response format of unique narrative replies allows for novel use cases, including acting as a simulated patient, a brainstorming tool providing individual feedback, and acting as a fellow classmate to simulate small-group style learning. For these applications to be useful, however, ChatGPT must exhibit a level of medical knowledge and reasoning that allows sufficient confidence in its responses.

In this paper we aim to assess the medical knowledge of ChatGPT by quantifying its performance through the use of two question sets centered around knowledge tested in the United States Medical Licensing Examination (USMLE) Step 1 and Step 2 exams. In addition, to further assess the ability of ChatGPT to serve as a virtual medical tutor, we qualitatively examine the integrity of ChatGPT’s responses with regards to logical justification and use of intrinsic and extrinsic information.

## METHODS

### Medical Education Datasets

We created two pairs of novel datasets to examine ChatGPT’s understanding of medical knowledge related to Step 1 and Step 2. We first selected a subset of 100 questions from AMBOSS, ^14^ a widely used question bank which contains over 2700 Step 1 and 3150 Step 2 CK questions. The existing performance statistics from previous AMBOSS users allows us to determine the models relative performance. We call these datasets AMBOSS-Step1 and AMBOSS-Step2, respectively. AMBOSS provides users with an Attending Tip when they have difficulty with a question, as well as a difficulty rating (1-5). We included these tips in our dataset to determine if additional context improves predictive performance.

We also used the list of 120 free Step 1 and Step 2 CK questions developed by the National Board of Medical Examiners (NBME), which we call NBME-Free-Step1 and NBME-Free-Step2, to evaluate ChatGPT’s performance on questions most closely aligned with those from the true licensure exams.

### Prompt Engineering

Due to the significant impact that prompt engineering has been shown to have on generative LM output, we standardized the input formats of the AMBOSS and NBME-Free datasets. First, we removed any questions that include an image, as ChatGPT only accepts textual input. Next we removed questions where the answer was formatted as a table. This was done so that the accuracy of ChatGPT’s answers was solely dependent on its ability to synthesize medical knowledge within narrative text, rather than parse complicated text inputs. Questions were formatted with the question text followed by the direct question separated by newline. In the AMBOSS datasets, the attending tip was inserted as a separate instance of the question. Following the question text and direct question, the multiple choice answers were provided, separated again by a newline. An example follows:

Example Question:

A 1-hour-old male newborn is examined in the hospital nursery. He was born at term to a 22-year-old woman, gravida 1, para 1, with HIV infection. The mother took antiretroviral therapy throughout her pregnancy, and her HIV viral load was consistently *<*1000 copies/mL. She received intravenous zidovudine (AZT) during labor. The patient’s Apgar scores were 8 and 9 at 1 and 5 minutes, respectively. He is at the 50th percentile for length, weight, and head circumference. He appears well. Examination shows no abnormalities.

In addition to administration of the hepatitis B vaccine, which of the following is the most appropriate pharmacotherapy for this newborn?

AMBOSS Attending Tip Here If Used

A. Lopinavir-ritonavir and trimethoprim-sulfamethoxazole
B. Lopinavir-ritonavir only
C. Penicillin
D. Trimethoprim-sulfamethoxazole only
E. Zidovudine (AZT)

Correct Answer:

(E)

ChatGPT Response:

The most appropriate pharmacotherapy for this newborn would be (B) Lopinavir-ritonavir only.

Lopinavir-ritonavir is a combination medication that is used as part of antiretroviral therapy (ART) to treat HIV infection. It is recommended for newborns exposed to HIV through mother-to-child transmission, such as in this case. The mother received antiretroviral therapy during her pregnancy, which likely reduced the risk of mother-to-child transmission of HIV. However, the newborn should still receive lopinavir-ritonavir as a prophylactic measure to further reduce the risk of HIV transmission and prevent the development of HIV infection.

Trimethoprim-sulfamethoxazole is a combination antibiotic that is used to treat infections caused by certain bacteria. It is not the most appropriate pharmacotherapy for this newborn because it is not used to treat or prevent HIV infection.

Penicillin is an antibiotic that is used to treat infections caused by certain bacteria. It is not the most appropriate pharmacotherapy for this newborn because it is not used to treat or prevent HIV infection.

Zidovudine (AZT) is a nucleoside reverse transcriptase inhibitor (NRTI) that is used as part of antiretroviral therapy (ART) to treat HIV infection. The mother received intravenous AZT during labor, which likely reduced the risk of mother-to-child transmission of HIV. However, the newborn should still receive lopinavir-ritonavir as a prophylactic measure to further reduce the risk of HIV transmission and prevent the development of HIV infection.

### Model Testing

We first recorded all correct answers as they appear in the AMBOSS and NBME datasets. We then prompted ChatGPT with the standardized questions, first without, then with the Attending Tip. Due to the nature of ChatGPT’s output, we manually reviewed each answer to determine which answer from the MCQ has been selected, if any.

We then qualified the ChatGPT responses for each question using three binary variables to assess:

1. Logical Reasoning: The response clearly identifies the logic in selecting between answers given information presented in the response.
2. Internal information: The response uses information internal to the question, including about the question in the response.
3. External information: The response uses information external to the question, including but not limited to qualifying the answers given or the stem.

Finally for each question answered incorrectly, we labeled the reason for the incorrect answer as one of four options:

- Logical Error: The response adequately found the pertinent information but did not properly convert the information to an answer.
  - Identifies young woman has having difficulty with taking pills routinely and still recommends oral contraceptives over an IUD.
- Information Error: ChatGPT either did not identify in a key piece of information, whether present in the question stem or external that would be considered expected knowledge.
  - Recommends antibiotics for sinusitis infection believing most cases to be of bacterial etiology even when the majority are viral.
- Statistical Error: An error centered around an arithmetic mistake. This includes explicit errors, such as stating “1 + 1 = 3” or indirect errors, such as an incorrect estimation of disease prevalence.
  - Identifies underlying nephrolithiasis, but misclassified the prevalence of different stone types.
- Multiple Errors: Any combination of the previous three options.

## RESULTS

From Table 1, ChatGPT performed more accurately on Step 1 related questions compared to Step 2 on both the NBME and AMBOSS datasets, 64.4% to 57.8% and 44% to 42% respectively. Similarly for both Step 1 and Step 2 questions, the model performed better on NBME questions when compared to AMBOSS, 64.4% to 44% and 57.8% to 42% respectively.

**Table 1.** Performance of ChatGPT on the four outline datasets.

|  | NBME-Free-Step1 | NBME-Free-Step2 | AMBOSS-Step1 | AMBOSS-Step2 |
|---|---|---|---|---|
| Number of Questions | 87 | 102 | 100 | 100 |
| Correct, n (%) | 56 (64.4) | 59 (57.8) | 44 (44.0) | 42 (42.0) |
| Incorrect, n (%) | 31 (35.6) | 43 (42.2) | 56 (56.0) | 58 (58.0) |

From Table 2, relative to AMBOSS users as reported on the after-test summary, ChatGPT was in the 30th percentile on Step 1 questions without the attending tip. On Step 1 with attending tip, as well as Step 2 with and without the attending tip the model performed at the 66th, 20th, and 48th percentiles respectively. On Step 1 questions without the attending tip ChatGPT had a significant decrease in accuracy as the AMBOSS reported difficulty increased (P=.012), falling from 64.3% accuracy on level 1 questions to 0.0% accuracy on level 5 questions. The remaining groups were monotonically decreasing in accuracy as question difficulty increased except for questions with difficulty 2 vs 3 for Step 1 with attending tip and questions with difficulty 4 and 5 for Step 1 without attending tip.

**Table 2.**
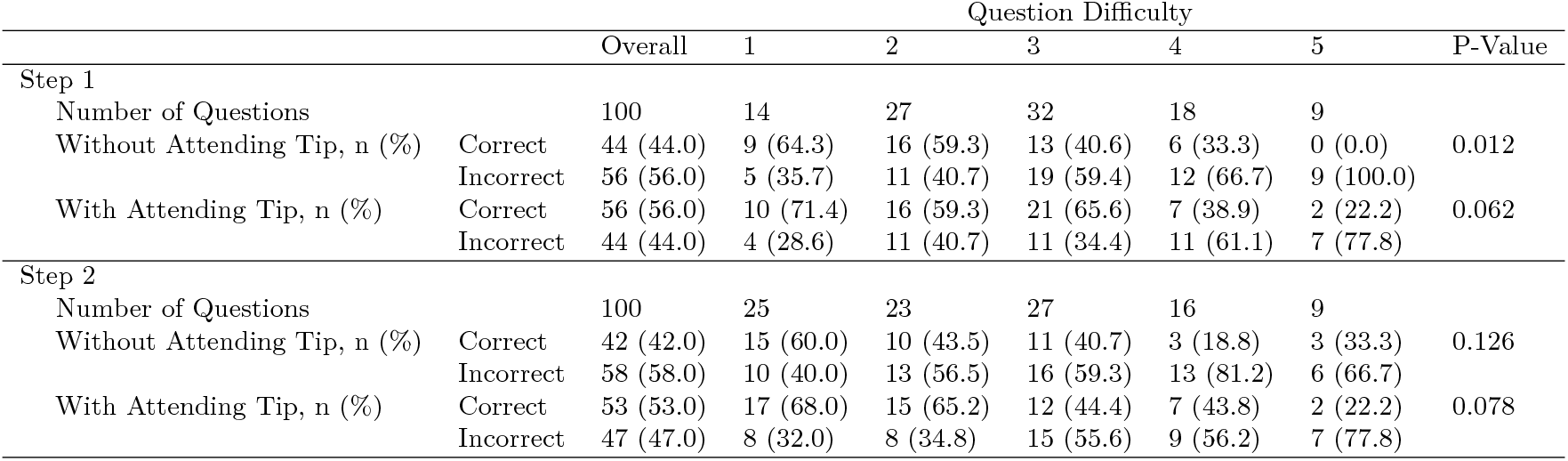
Model Performance on AMBOSS-Step1 and AMBOSS-Step2 datasets by question difficulty.

|  |  | Question Difficulty |  |  |  |  |  |  |
|  |  | Overall | 1 | 2 | 3 | 4 | 5 | P-Value |
|---|---|---|---|---|---|---|---|---|
| Step 1 |  |  |  |  |  |  |  |  |
| Number of Questions |  | 100 | 14 | 27 | 32 | 18 | 9 |  |
| Without Attending Tip, n (%) | Correct | 44 (44.0) | 9 (64.3) | 16 (59.3) | 13 (40.6) | 6 (33.3) | 0 (0.0) | 0.012 |
|  | Incorrect | 56 (56.0) | 5 (35.7) | 11 (40.7) | 19 (59.4) | 12 (66.7) | 9 (100.0) |  |
| With Attending Tip, n (%) | Correct | 56 (56.0) | 10 (71.4) | 16 (59.3) | 21 (65.6) | 7 (38.9) | 2 (22.2) | 0.062 |
|  | Incorrect | 44 (44.0) | 4 (28.6) | 11 (40.7) | 11 (34.4) | 11 (61.1) | 7 (77.8) |  |
| Step 2 |  |  |  |  |  |  |  |  |
| Number of Questions |  | 100 | 25 | 23 | 27 | 16 | 9 |  |
| Without Attending Tip, n (%) | Correct | 42 (42.0) | 15 (60.0) | 10 (43.5) | 11 (40.7) | 3 (18.8) | 3 (33.3) | 0.126 |
|  | Incorrect | 58 (58.0) | 10 (40.0) | 13 (56.5) | 16 (59.3) | 13 (81.2) | 6 (66.7) |  |
| With Attending Tip, n (%) | Correct | 53 (53.0) | 17 (68.0) | 15 (65.2) | 12 (44.4) | 7 (43.8) | 2 (22.2) | 0.078 |
|  | Incorrect | 47 (47.0) | 8 (32.0) | 8 (34.8) | 15 (55.6) | 9 (56.2) | 7 (77.8) |  |

Finally, in Table 3, we evaluated ChatGPT’s answer quality across three metrics as outlined above, presence of logic, internal information, and external information. We found that every response provided by ChatGPT provided a logical explanation of its answer selection, independent of the correctness of the response. Additionally, across both NBME-Free-Step1 and NBME-Free-Step2 datasets, for both correct and incorrect responses, ChatGPT used information internal to the question in greater than 90% of questions. There was no significant difference between presence of internal information between correct or incorrect responses for either Step 1 or Step 2 datasets (P=.25 and .071). Finally, information external to the question was used in 92.9% of correct responses and 42.4% of incorrect for the Step 1 dataset (P*<*.001). For the Step 2 dataset external information was used in and 89.8% of correct and 62.8% of incorrect answers.(P=.001). For both step 1 and step 2 logical errors were the most common, followed by information errors. Few statistical errors were present for either dataset.

**Table 3.** Qualitative analysis of answer quality for NBME-Free-Step1 and NBME-Free-Step2 datasets

|  |  | NBME-Free-Step1 |  |  | NBME-Free-Step2 |  |  |
|---|---|---|---|---|---|---|---|
|  |  | Overall | Correct | Incorrect | Overall | Correct | Incorrect |
| Number of Quesions, n |  | 87 | 56 | 31 | 102 | 59 | 43 |
| Logical Reasoning, n (%) | True | 87 (100.0) | 56 (100.0) | 31 (100.0) | 102 (100.0) | 59 (100.0) | 43 (100.0) |
|  | False | 0 (0.0) | 0 (0.0) | 0 (0.0) | 0 (0.0) | 0 (0.0) | 0 (0.0) |
| Internal Information, n (%) | True | 84 (96.6) | 55 (98.2) | 29 (93.5) | 99 (97.1) | 59 (100.0) | 40 (93.0) |
|  | False | 3 (3.4) | 1 (1.8) | 2 (6.5) | 3 (2.9) | 0 (0.0) | 3 (7.0) |
| External Information, n (%) | True | 67 (77.0) | 52 (92.9) | 15 (48.4) | 80 (78.4) | 53 (89.8) | 27 (62.8) |
|  | False | 20 (23.0) | 4 (7.1) | 16 (51.6) | 22 (21.6) | 6 (10.2) | 16 (37.2) |
| Reason for Incorrect Answer, n (%) | Logical Error |  | 13 (41.9) |  |  | 16 (37.2) |  |
|  | Information Error |  | 7 (22.6) |  |  | 13 (30.2) |  |
|  | Statistical Error |  | 2 (6.5) |  |  | 1 (2.3) |  |
|  | Multiple Errors |  | 9 (29.0) |  |  | 13 (30.2) |  |

## DISCUSSION

ChatGPT marks a significant advancement in the field of NLP. ^9^ One of the key features touted by ChatGPT is its ability to understand context and carry on a conversation that is coherent and relevant to the topic at hand. In this paper we have shown that this extends into the medical domain by evaluating ChatGPT on four unique medical QA datasets. We found that the model is capable of correctly answering up to over 60% of questions representing topics covered in the USMLE Step 1 and Step 2 licensing exams. A threshold of 60% is often considered the benchmark passing standards for both Step 1 and Step 2, indicating that ChatGPT performs at the level expected of a 3rd year medical student. Additionally, our results demonstrate that even in the case of incorrect answers, the responses provided by the model always contain a logical explanation for the answer selection, and greater than 90% of the time this response directly includes information contained in the question stem. Correct answers were found to contain information external to the question stem significantly more frequently than incorrect responses, indicating that the ability of the model to correctly answer a question may be related to its ability to relate the prompt to data within its corpus.

Prior work in Medical Question and Answer research has often been focused on more specific tasks with the intent of improving model performance at the expense of generalizability. For example, Jin et al. achieved a 68.1% with their model which answers Yes/No questions who’s answers may be found in the corpus of Pubmed available abstracts. ^15^ Attempts at more generalizable models have been met with more challenges. A different Jin et al. achieved an accuracy of 36.7% on a dataset of 12,723 questions derived from Chinese medical licensing exams. ^16^ Similarly, in 2019 Ha et al. reported only a 29% acuraccy on 454 USMLE Step 1 and Step 2 questions. ^17^ ChatGPT therefore thus represents a significant step forward on three distinct fronts. First is generalizability, as ChatGPT is capable of responding to any question which can be formatted with text alone; the scope of possible questions is limited only by what can be submitted by the user. Second is accuracy. We have shown that ChatGPT equals or outperforms prior models on questions of similar difficulty and content. Finally, ChatGPT marks the greatest jump forward in user interpretability due to its conversational interface. Each response has some level of reasoning as we have demonstrated. And the ability to ask follow up questions allows the user to gain a larger perspective on the concept being addressed in the question, rather than just an answer output alone.

One potential use case for the model is as an adjunct or surogate for small group education. Small group education has been shown to be a highly efficacious method of teaching. ^18^ Not to mention the value of the small group amidst problem-based learning in medicine. ^19^ Other examples facilitating small group discourse in medical education include clinical problem solving by working through case presentations. ^20^ The education is useful independent of the knowledge of the students, as evidenced by small group based learning starting as early as the first week after matriculation. What is often beneficial is the ability of students to test ideas off of each other and receive feedback. With its dialogic interface, ChatGPT is able to provide these same benefits for students when they are studying independently. Students could use the tool to ask questions about specific medical concepts, diagnoses, or treatments, and receive accurate and personalized responses to help them better structure their knowledge around each concept. As the technology is further explored and improved, it is also possible that novel educational methods may be developed which fully utilize the capabilities of a tool like ChatGPT.

## CONCLUSION

In conclusion, we have demonstrated that ChatGPT has an amount of medical knowledge to allow for its use as a novel educational tool. It performs at the level of a third year medical student on the question sets examined, and its responses to questions provide interpretable context to justify models written response in most cases. By providing personalized, easily-interpretable responses, the model allows for the creating of an on-demand, interactive learning environment for students which can allow for increased information retention and a more enjoyable learning experience.

The conclusion to the paper was written by ChatGPT from the prompt “Write a conclusion of the following manuscript:” with the entire text of the manuscript included afterwards:

In this manuscript, the authors evaluated the medical knowledge of ChatGPT, a natural language processing (NLP) model, using two sets of questions related to the USMLE Step 1 and Step 2 exams. They created two pairs of datasets from AMBOSS and the National Board of Medical Examiners (NBME) and standardized the input format of the questions to remove any image or table-based answers. The authors then used these datasets to assess ChatGPT’s performance on medical knowledge and determined its relative performance by comparing it to existing performance statistics from AMBOSS users. The results of this study suggest that ChatGPT has some understanding of medical knowledge, but further research is needed to determine its usefulness as a tool for medical student learning and other applications in the medical field.

## Data Availability

All data produced in the present work are available online at the following link:

https://bit.ly/3FTLfJ0

## ACKNOWLEDGMENTS

This publication was made possible by the Yale School of Medicine Fellowship for Medical Student Research. Research reported in this publication was supported by the National Institute of Diabetes And Digestive And Kidney Diseases of the National Institutes of Health under Award Number T35DK104689. The content is solely the responsibility of the authors and does not necessarily represent the official views of the National Institutes of Health.

## CONFLICTS OF INTEREST

All authors listed certify that they have no affiliation of involvement in any organization with financial or non-financial interest in the subject matter of this manuscript.

### ABBREVIATIONS

NLP: Natural Language Processing
NBME: National Board of Medical Examiners
USMLE: United States Medical Licensing Examination
QA: Question-Answering
AZT: Zidovudine (AZT)
NRTI: Nucleoside reverse transcriptase inhibitor
ART: Antiretroviral therapy

## SUPPLEMENTARY INFO

All questions, annotations, and ChatGPT responses can be found here: https://bit.ly/3FTLfJ0

## REFERENCES

[1] Scott Kevin. Microsoft teams up with openai to exclusively license GPT-3 language model 2020.

[2] Bowman Emma. A new AI chatbot might do your homework for you. but it’s still not an A+ student 2022.

[3] How good is chatgpt? https://www.economist.com/business/2022/12/08/how-good-is-chatgpt2022.

[4] Can Artificial Intelligence (chat GPT) get a 7 on an SL maths paper? 2022.

[5] Das Avisha, Selek Salih, Warner Alia R., et al. Conversational Bots for Psychotherapy: A Study of Generative Transformer Models Using Domain-specific Dialogues in Proceedings of the 21st Workshop on Biomedical Language Processing(Dublin, Ireland):285– 297Association for Computational Linguistics 2022.

[6] Savery Max, Abacha Asma Ben, Gayen Soumya, Demner-Fushman Dina. Question-driven summarization of answers to consumer health questions Scientific Data. 2020;7:1–9.

[7] Logé Cécile, Ross Emily, Dadey David Yaw Amoah, et al. Q-Pain: A Question Answering Dataset to Measure Social Bias in Pain Management arXiv preprint 2108.01764. 2021.

[8] Gutiérrez Bernal Jiménez, McNeal Nikolas, Washington Clay, et al. Thinking about GPT-3 In-Context Learning for Biomedical IE? Think Again arXiv preprint 2203.08410. 2022.

[9] Schulman John, Zoph Barret, Kim Christina, et al. ChatGPT: Optimizing Language Models for Dialogue 2022.

[10] Google Scholar https://scholar.google.com/ 2022.

[11] UpToDate https://www.uptodate.com 2022.

[12] DynaMed https://www.dynamed.com/ 2022.

[13] PubMed https://pubmed.ncbi.nlm.nih.gov/ 2022.

[14] Medical knowledge platform for doctors and students

[15] Jin Qiao, Dhingra Bhuwan, Liu Zhengping, Cohen William W, Lu Xinghua. PubMedQA: A dataset for biomedical research question answering arXiv preprint 1909.06146. 2019.

[16] Jin Di, Pan Eileen, Oufattole Nassim, Weng Wei-Hung, Fang Hanyi, Szolovits Peter. What Disease Does This Patient Have? A Large-Scale Open Domain Question Answering Dataset from Medical Exams Applied Sciences. 2021;11:6421.

[17] Ha Le An, Yaneva Viktoriya. Automatic question answering for medical MCQs: Can it go further than information retrieval? RANLP 2019.

[18] Springer Leonard, Stanne Mary Elizabeth, Donovan Samuel S.. Effects of Small-Group Learning on Undergraduates in Science, Mathematics, Engineering, and Technology: A Meta-Analysis Review of Educational Research. 1999;69:21–51.

[19] Neville Alan J, Norman Geoff R. PBL in the undergraduate MD program at McMaster University: three iterations in three decades Academic Medicine. 2007;82:370–374.

[20] Anspach Renee R. Notes on the sociology of medical discourse: The language of case presentation Journal of health and social behavior. 1988:357–375.

